# Model Based Covid-19 Case Studies in the UK, the USA and India

**DOI:** 10.1101/2020.05.31.20118760

**Authors:** Santanu Basu

## Abstract

Time dependent spread of Covid-19 among the population of the UK, the USA and India is analyzed using a recently developed mathematical model [1-3]. Results of model predictions of case growth in these countries during the next six weeks are also presented. The model is applicable to case studies and near term predictions for other countries and regions.

## INTRODUCTION

Almost all countries in the world have severely suffered from the rapid spread of the Covid-19 disease caused by the new coronavirus, SARS-CoV-2 during the first few months of 2020. A few things are in common for all countries:

- The virus is thought to spread mainly from person to person [4]. The virus seems to enter a healthy person’s body through the mouth or the nose when the person is in proximity of an infected person (about 6 ft or less) or when the healthy person touches her or his own face after touching a virus laden surface.
- The number of people with symptoms is less than 10 at the onset of the outbreak of this disease in each country.
- Testing for the disease was developed in the Jan 20^th^ - Feb 15^th^ time frame. The total number of Covid-19 cases in the world on Feb 15^th^ was 69,000 (1.2% of the current total).
- The vaccine for this disease is not available in any country as of May 31^st^.

Despite having these attributes in common, the growth of the number of cases and the casualties differed greatly from country to country. For example the number of cases per million inhabitants in Spain is 19 times than that of New Zealand and the number of deaths per million inhabitants in Spain is 131 times than that of New Zealand as of May 25^th^ 2020. The total number of casualties is thus a strong function of the person to person separation achieved during the outbreak in each country which mostly was governed by the social distancing, quarantine and patient isolation measures taken by the authorities in each country or region within a country. The steps taken or not taken by the authorities did make a big difference. The objective of this paper is to study the effect of the disease transmission parameters in a quantitative manner on case results in various countries.

Mathematical modeling of spreading of infectious diseases is a well-established field [5-6]. There have been a large number of recent efforts in modelling Coronavirus spreading as reported in the literature [7-9]. A mathematical model has been recently developed that can efficiently model the Covid-19 case growth over a large dynamic range of casualty rate [1-3]. In this paper we will apply this model to study the growth of Covid-19 cases in the UK, the USA and India with 23% of the world’s population and make prediction for the growth of cases during the next six weeks.

## OVERVIEW OF THE COVID-19 MATHEMATICAL MODEL

We developed and documented a mathematical model to predict number of cases and hospital patient loads [1-3]. The model is included in the appendix. The model assumes that starting from a small number of infected people which act a seed, the infection grows by transmission from person to person in proximity. The model keeps track of the number of infected people at various stages of development of the disease through the course of the virus outbreak. The model is parametric and transparent so that predictions are based on parameters which are in principle measurable and the predictions can be continuously corrected as more accurate data on parameters become available. Typical machine learning and curve fitting models which are not based on any real-life parameters tend to have high degree of error in making projections and do not provide physical insight into the problem that can be used in the future to minimize casualties. At present, the model has twenty one user-supplied variable parameters, three of which influence the results greatly. These three are (1) n_1_, the first date of implementation of social distancing protocols by the authorities since the viral outbreak, (2) 1/G, the rate at which the transmission decreases during implementation of the measures and (3) T_f_, the final rate of transmission in the long run during the first outbreak.

The objective of this paper is to analyze the cases of three large countries in terms of population. At present lifting of stay at home orders are under way in these three countries which should have an effect on the number of cases trend. Therefore we limit our prediction to only six weeks in the future. It is hoped that comparing three countries in three continents with the same mathematical model should reveal some underlying pattern which can be a valuable lesson to minimize the spread of an infectious disease and loss of lives for now and in the future.

## RESULTS AND DISCUSSION

### The UK

Covid-19 case data for the UK was obtained from [10-11]. The first case of Covid-19 in the UK was confirmed on January 31^st^, 2020. The authorities started taking measures to isolate the symptomatic patients on March 12^th^ and stay at home orders were finally issued on March 23^rd^. The number of confirmed Covid-19 cases grew from 1,579 to 11,084 during this period.

The UK data could be fitted well with March 22^nd^ to be the first date (n_1_) of transmission rate decrease, 9 days for exponential time constant for transmission rate decrease (G) and 4.5% for the final transmission rate (T_f_). The comparison between the model predictions until July 31^st^ and actual data until June 8^th^ on number of cases and deaths are shown in figure 1. The figures demonstrate that the model fits the actual data very well.

**Figure 1.**
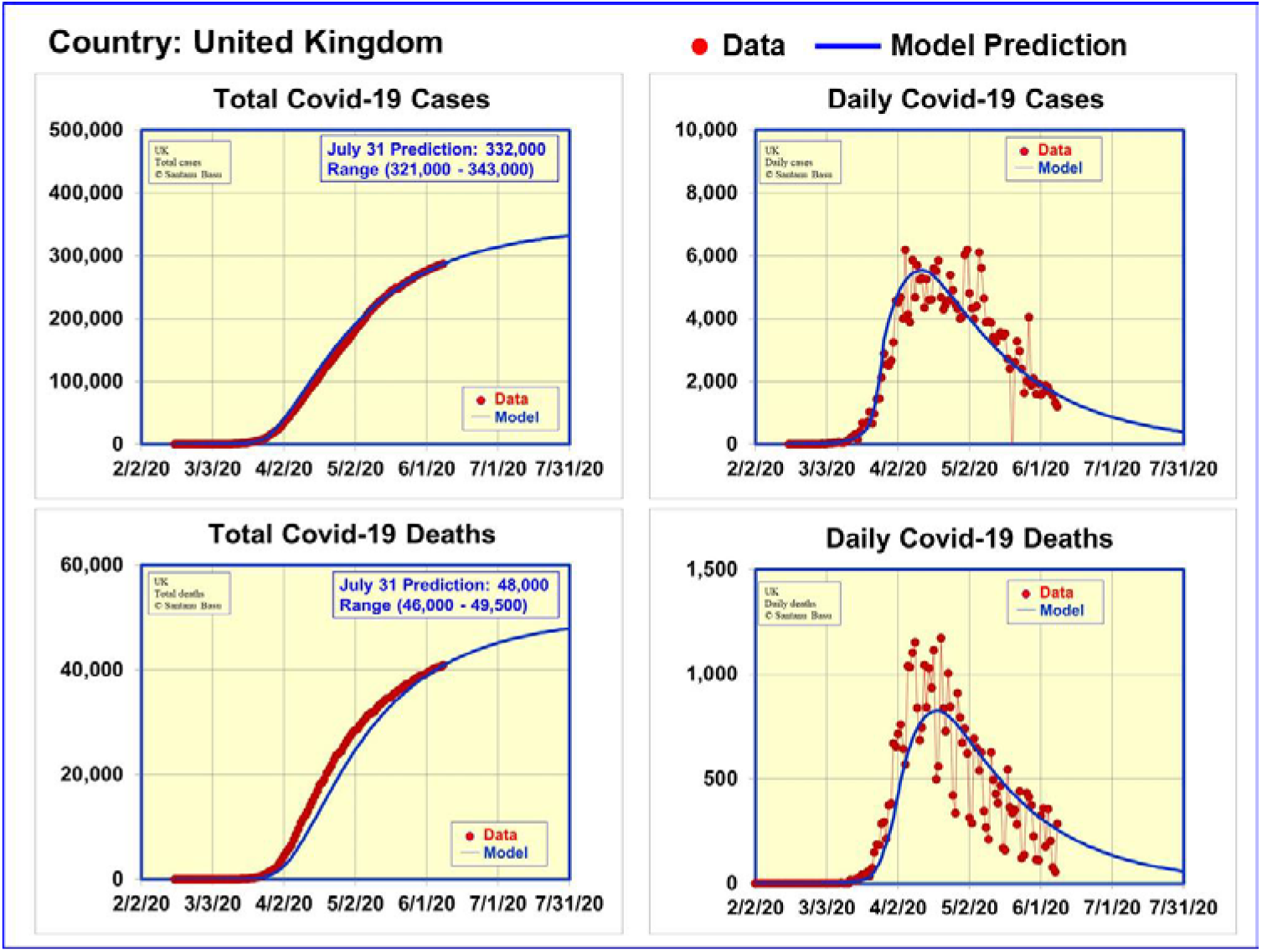
Data and model predictions for Covid-19 cases and deaths in the UK

Having anchored the model to actual data until June 8^th^, we are in a position to make reliable projections for six weeks in advance. The model predicts the number of cases to rise to 332,000 (range 321,000-343,000) and the number of deaths to rise to 48,000 (range 46,000-49,500) on July 31^st^, 2020. The model also predicts the daily number of cases to fall to 400 (range 250-560) and the number of daily deaths to fall to 60 (range 40-80) on the same date.

### The USA

We obtained the case data for USA from [12-13]. The first confirmed case of Covid-19 was on January 20^th^, 2020 [13]. The authorities declared a public health emergency on January 31^st^. Stay at home orders were issued in several counties in California on March 17^th^ and the Surgeon General endorsed physical distancing measures on March 23^rd^. The stay at home practice differed from state to state and was in place nearly everywhere by April 11^th^ [13]. The numbers of Covid-19 cases with symptoms in the USA were 4,661 on March 17^th^ and 501,560 on April 11^th^ [12].

The USA data could be fitted well with March 16^th^ to be the first date (n_1_) of transmission rate decrease, 9 days for exponential time constant for transmission rate decrease (G) and 5.2% for the final transmission rate (T_f_). The comparison between the model predictions until July 31^st^ and actual data until June 8^th^ on number of cases and deaths are shown in figure 2. The figures demonstrate that the model fits the actual data for the USA very well.

**Figure 2.**
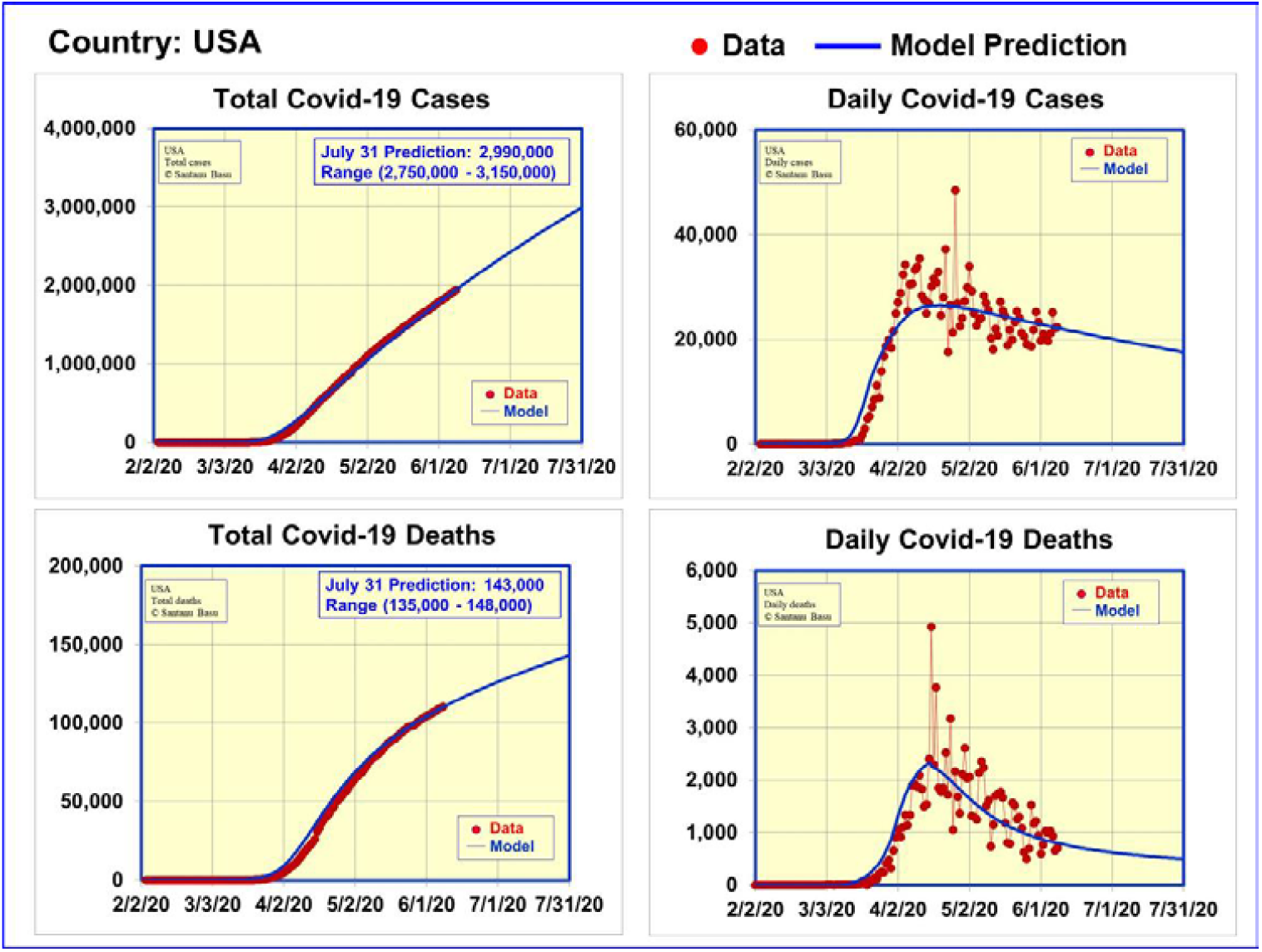
Data and model predictions for Covid-19 cases and deaths in the USA

With the model anchored to actual data until June 8^th^, reliable predictions can be made for the expected number of cases and deaths for six weeks in advance. The model predicts the number of cases to rise to 2.99 million (range 2.75 to 3.15 million) and the number of deaths to rise to 143,000 (range 135,000–148,000) on July 31^st^, 2020. The model also predicts the daily number of cases to fall to 17,500 (range 12,000-21,000) and the number of daily deaths to fall to 500 (range 340-600) on the same date.

### India

We obtained the case data for India from [14]. The first confirmed case of Covid-19 was on January 30^th^, 2020 [14]. As in almost all countries, the Covid-19 in India started with people coming from abroad. Protective measures to screen selected airport passengers started on January 21^st^ and screening of all international air travelers began on March 8^th^. Schools started closing on March 6^th^ and various forms of social distancing measures were being adopted at different parts of the country. Finally a nationwide lockdown was announced on March 24^th^. The number of Covid-19 cases with symptoms in India was 31 on March 6^th^ which grew to 519 on March 24^th^ [14]. The case count in India was lower when the authorities took action to limit the spread of Covid-19 than in the UK or the USA which should have a positive effect. However the challenge for India is very high population density (1,087 per sq mi in India as compared to 725 per sq mi in the UK and 87 per sq mi in the USA) which makes keeping 6 ft of person to person separation much harder. The long term transmission rate (T_f_) is expected to be higher in India.

India Covid-19 case data could be fitted well with March 22^nd^ to be the first date (n_1_) of transmission rate decrease, 42 days for exponential time constant for transmission rate decrease (G) and 5.2% for the final transmission rate (T_f_). The comparison between the model predictions until July 15^th^ and actual data until June 8^th^ on number of cases and deaths are shown in figure 3. The figures demonstrate that the model fits the actual data for India very well.

**Figure 3.**
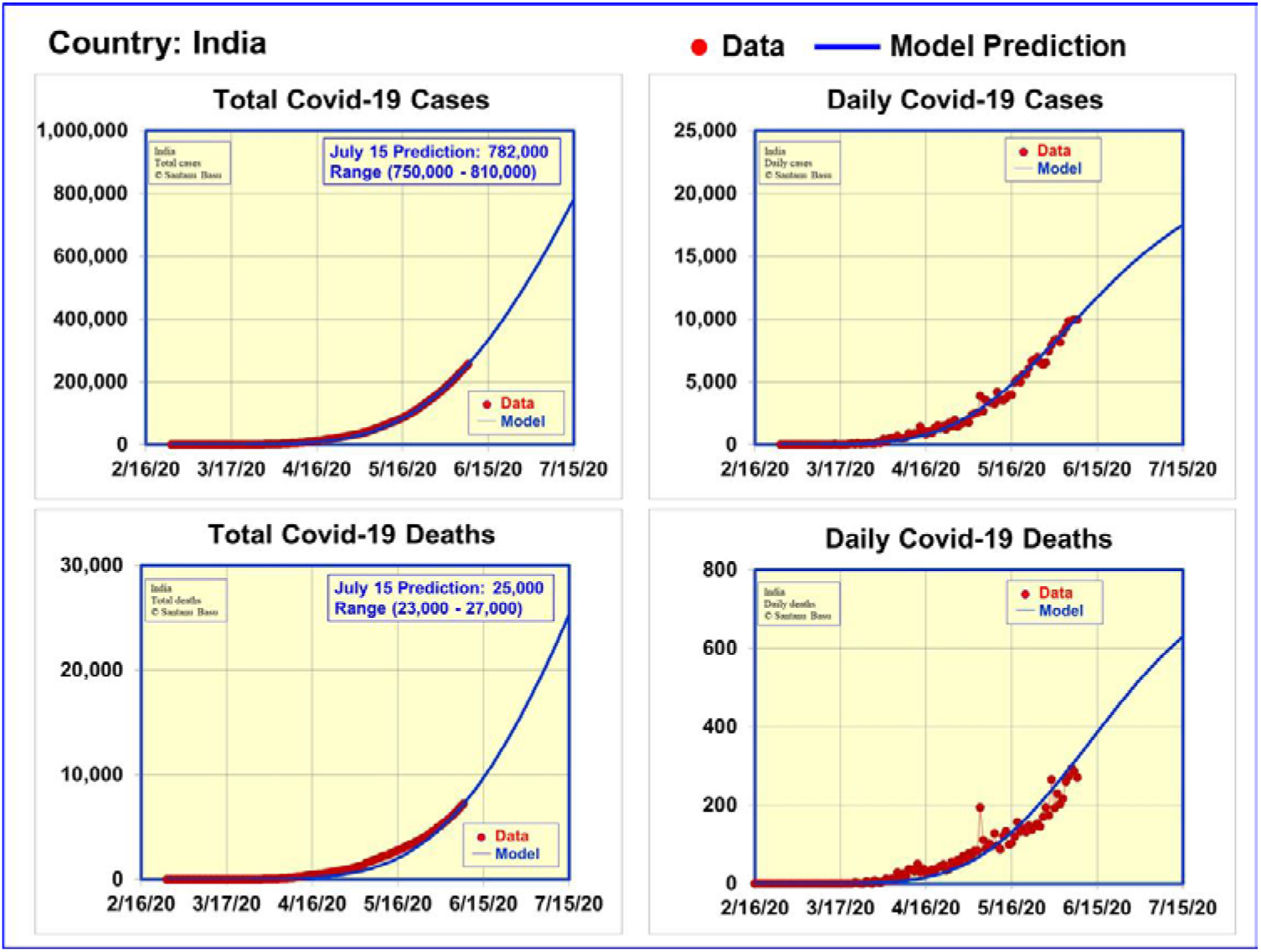
Data and model predictions for Covid-19 cases and deaths in India

Having the model anchored to actual data until June 8^th^, predictions are made for the growth of number of cases and deaths. The difference between India and the UK and the USA is that the onset of the disease is a little later in India and the daily case numbers are still increasing. We limited the prediction to July 15^th^ for India. The number of cases is predicted to rise to 782,000 (range 750,000-810,000) and the number of deaths to rise to 25,000 (range 23,000-27,000) on July 15^th^, 2020. The model also predicts the daily number of cases to rise to 17,500 (range 16,000-19,000) and the number of daily deaths to rise to 630 (range 560-700) on the same date.

Figure 4 shows the number of hospital beds needed for Covid-19 patients in India as a function of time until July 15^th^. Reliable data on the number of Covid-19 patients who have been hospitalized in India as a function of time to date is not available. Therefore we could not anchor our model to available data. In spite of that, we assumed 16% of the Covid-19 patients are requiring hospitalization, out of whom 35% are requiring ICU beds to estimate the hospital bed requirements for India. The results are shown in figure 4. The model predicts the required number of hospital beds to be 22,000 (range 20,000-24,000), 35% of which need to be ICU beds.

**Figure 4.**
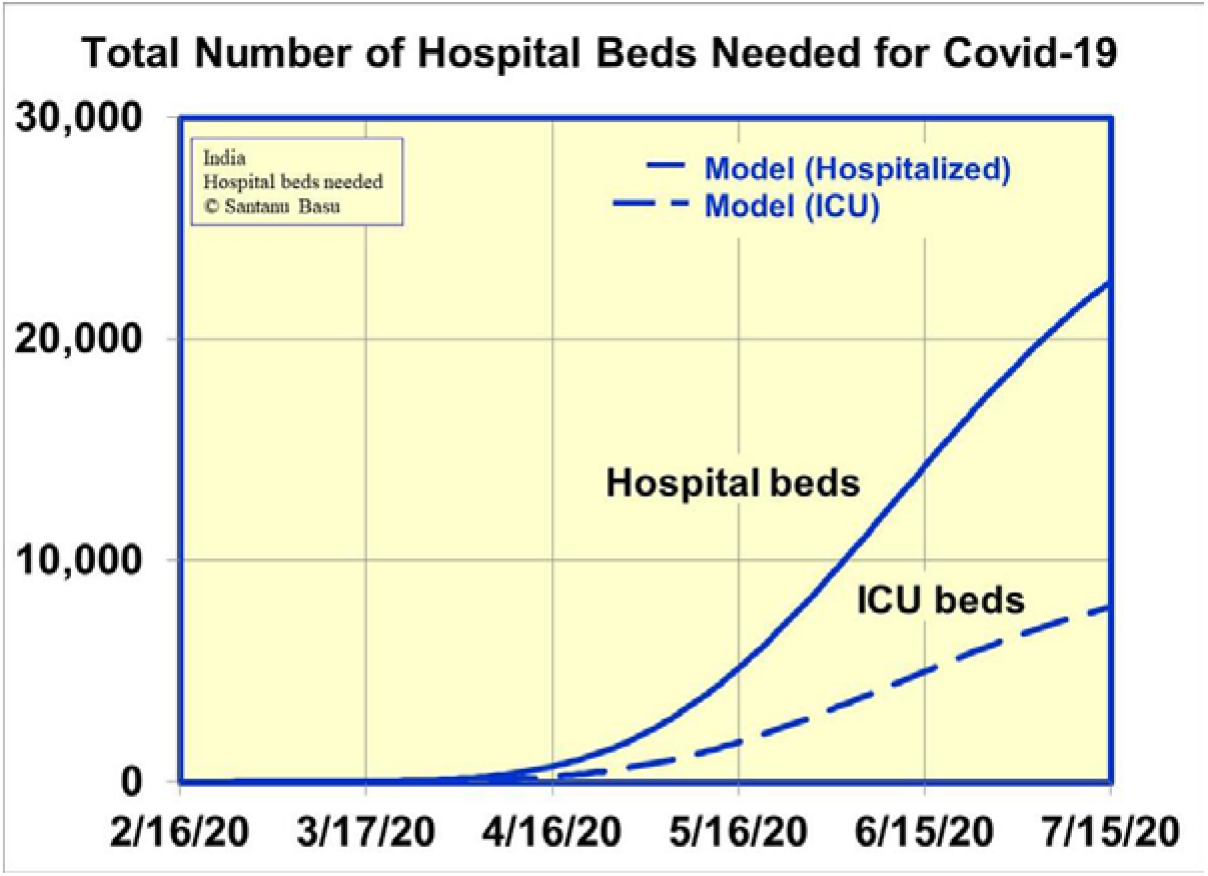
Model predictions for hospital bed requirements for Covid-19 patients in India

In conclusion, we have developed a mathematical model that provides insight into the Covid-19 viral outbreak cases in any country. The model fits the data very well for the UK, the USA and India with different levels of cases and casualties. We also presented predictions from the model for the number of cases six weeks in advance which can be very valuable for providing medical services to reduce casualties. The prediction for India has been found to be good after comparing with actual data [26]. The model is transparent and is based on parameters which are measurable. This allows the model calculations to be updated as the actual data of the parameters become available. Overall the model makes a hopeful conclusion that a great number of human lives can be saved by reducing the three parameters n_1_, G and T_f_ in the model which in everyday parlance means by taking timely social distancing and patient isolation measures. Timeliness of doing the necessary but difficult action of keeping people separated by about 6 ft is the key to success in this case before ultimately a vaccine becomes available.

The author wishes to acknowledge valuable discussions with P. Soni, L. Gutheinz, N. Shah, P. Hagelstein, W. Grossman, S. Gowrinathan, S. Watanabe, J. Basu, T. Hausken, S. Sheng, D. Tauber, T. Mitra, S. Dhawan, M. Genevro and J. Albertine.

## Data Availability

The data is available in the paper and in the websites linked to the document

https://coronavirus.data.gov.uk/#category=nations&map=rate&area=e92000001

https://ourworldindata.org/covid-cases

https://en.wikipedia.org/wiki/COVID-19_pandemic_in_India

## Additional Information

Correspondence and requests for materials should be addressed to the author.

## APPENDIX

In this section we study the propagation of a new infectious disease through an isolated country’s population, P. With some simple modifications, the procedure can be applied to recurring outbreak of the same virus and also to the normal state in which there is population movement among countries.

Figure 5 is a schematic of our model. Let n be the number of days since the onset of virus infection. The entire population of virus infected people is classified in six categories:

**Figure 5.**
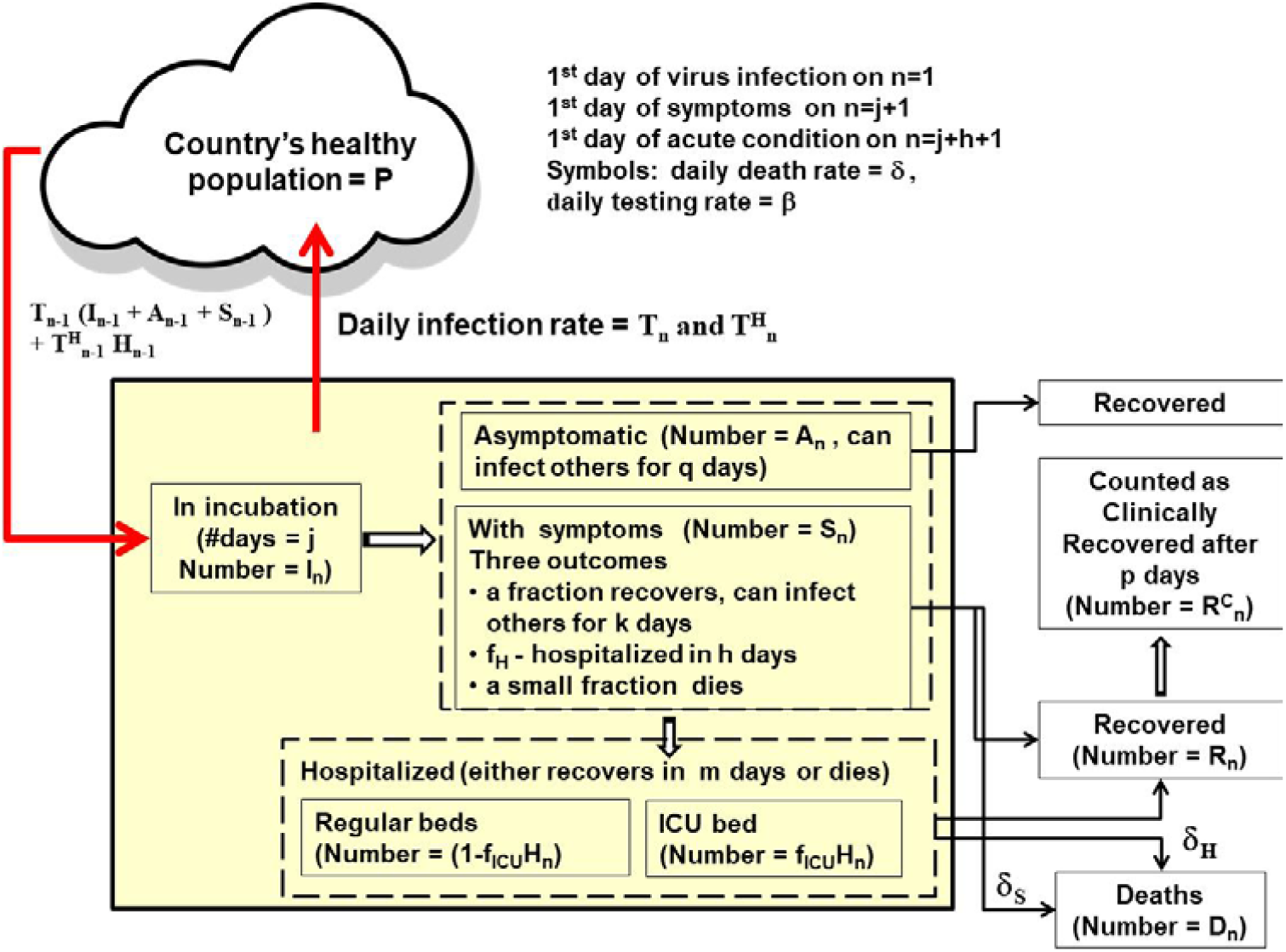
Schematic of the mathematical model for the virus infected cases

I – people in whom the virus is in incubation

A – people who are infected with the virus but who do not exhibit any symptoms

S – non-hospitalized patients who exhibit mild symptoms of the viral infection

H – hospitalized patients who are in acute condition

D – people who have died from the disease

R – people who have recovered from the illness and awaiting to be declared clinically recovered R^C^ – people who are counted as clinically recovered

On any day n, the total number of people who have been infected with the virus is N_n_, where

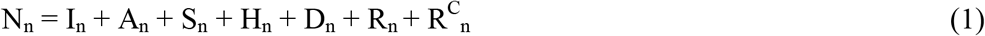

We denote the incubation period for the virus in number of days by j. The model assumes each infected person spends j days in incubation and is capable of infecting others. Let I_n_ be the total population of infected people who are still in the incubation period on day n. Let I n ^i^ be the population of those infected people in the incubation period on day n who have been infected for i number of days (i ranges from 1 to j).

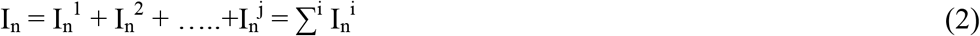

We assume that after the incubation period of j days, each infected person either is asymptomatic or shows symptoms of the disease. Let A n ^i^ be the population on day n of those asymptomatic people who have been carrying the virus for j+i number of days (1 ≤ i ≤ q) and let A_n_ be the total population of asymptomatic people who are capable of infecting others.

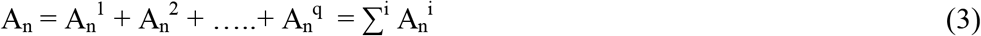

Let S _n_^i^ be the population on day n of non-hospitalized patients who have been showing mild symptoms for i number of days (1 ≤ i ≤ k) and let S_n_ be the total population of non-hospitalized patients.

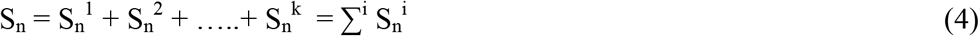

For each person with symptoms, there are three outcomes. A fraction, f_H_ is hospitalized, a small fraction dies at a rate of δ_S_ per person per day, and the remaining fraction recovers. The average time between showing symptoms and hospitalization is h ≤ k. We assume each patient with symptoms is capable of infecting others for a total of j+k number of days.

Let H_n_ be the total population of patients who are hospitalized (in acute condition) on day n. Let H _n_^i^ be the population on day n of those infected patients who have been hospitalized for i number of days (i ranges from (1 ≤ i ≤ m).

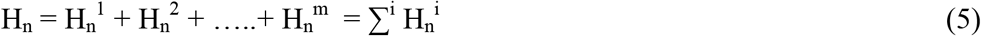

Out of the hospitalized patients, a fraction f_ICU_ is admitted to the ICU, the remaining fraction (1-f_ICU_) requires a regular hospital bed. The number of patients in the ICU is

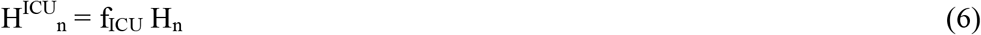

We denote the probability of death among the hospitalized patients to be δ_H_^i^ per day and the length of hospital stay to be m number of days. The patients who survive m days in the hospital are recovered but generally needs to wait p more days before being counted as clinically recovered.

The total number of people who are currently infected on day n and who are able to transmit the virus is (I_n_ + A_n_ + S_n_ +H_n_). Out of this population, only (S_n_ + H_n_) number of people show outward signs of the disease. Ironically in the beginning of the outbreak, when (S_n_ + H_n_) is small, the problem is sometimes underestimated without realizing that the disease is spreading exponentially everyday by the entire infected population (I_n_ + A_n_ + S_n_ +H_n_). The ratio (I_n_ + A_n_ + S_n_ +H_n_) / (S_n_ + H_n_) can be very high at the onset of the viral outbreak. The time to take swift, decisive and uniform measures to prevent spreading of the disease is when (S_n_ + H_n_) is small (for example less than 10).

We denote the rate of transmission of the virus from an infected person to a healthy person per day by T_n_. Another way of quantifying the rate of transmission that has been used in the literature is infection reproductive number which for SARS-CoV-2 is reported to be ∼ 2.5 [15-16]. Let N_0_ be the number of infected people on the first day. The numbers of infected people on the first two days of the outbreak are:

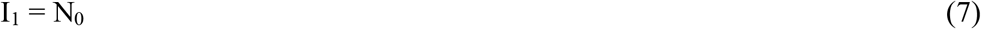

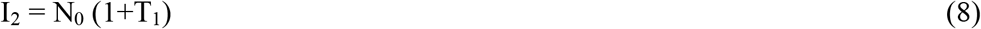

In general, the number of newly infected people on day n is given by

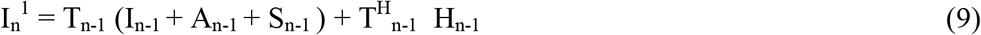

where T^H^_n_ is the rate of transmission from an hospitalized patient to a healthy person. In a well-managed hospital, T^H^_n_ = 0. A fraction, f_a_, of the newly infected people is asymptomatic of the disease. An estimate of f_a_ = 30% has been given in [17] which has been used in this model calculations. The number of new patients with mild symptoms on day n is given by

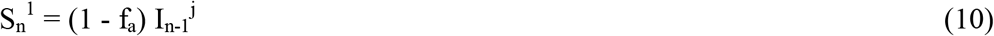

Similarly the number of new asymptomatic patients on day n is given by

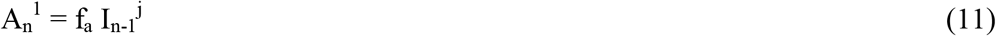

The number of new hospitalized patients on day n is given by

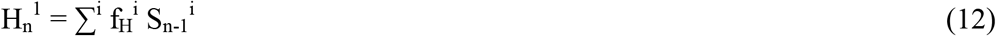

For simplicity, we can assume f_H_^i^ =0 for i ≠ h. The numbers of patients who die and recover from mild symptoms on day n are given by ∑^i^ δS ^i^ S_n_^i^ and (1-δ _S_^k^) S_n-1_^k^ respectively. Similarly the numbers of hospitalized patients who die and recover from acute conditions on day n are given by ∑^i^ δ_H_^i^ Hn ^i^ and (1-δ_H_^m^) H _n-1_^m^ respectively.

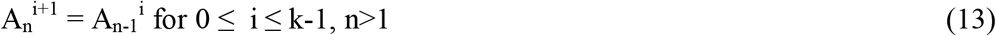

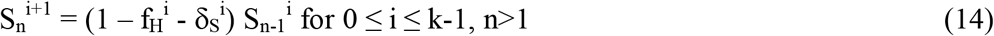

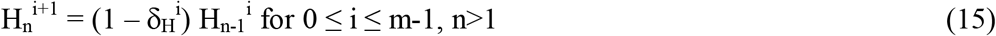

The number of deaths caused by the virus on day n is given by

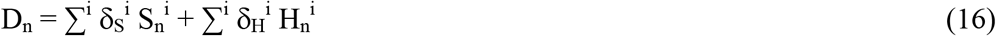

The number of patients who recover from the disease on day n is given by

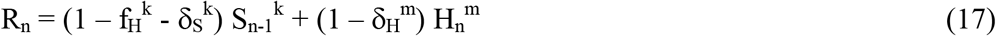

After an infected person first recovers either from the mild symptom state or after being hospitalized, it takes p number of days for her or him to be counted as clinically recovered which may vary from county to county and state to state. The number of new cases of clinically recovered ex-patients on day n is given by

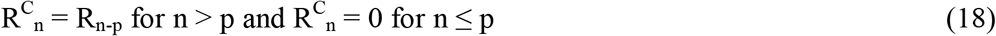

Before the first person showing symptoms of the disease caused by the new virus and shortly thereafter up to day n_1_, the transmission rate T_n_ is assumed to be constant at an initial value T_0_:

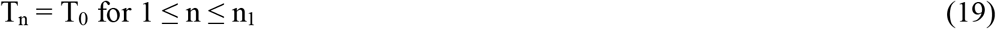

Typically the authorities take steps to prevent the spreading of the disease such as patient isolation and testing, contact tracing and social distancing. As a result T_n_ decreases with time. The model is capable of including actual time dependent transmission data. However at this time such data for the Coronavirus case is not available. A reasonable approximation is exponential decay of the rate of transmission T_n_ from infected persons to the general population. Let G be the number of days it takes for T_n_ - T_f_ to decrease by 63% of T_0_ - T_f_, and let T_f_ be the long term steady state value of T_n_.

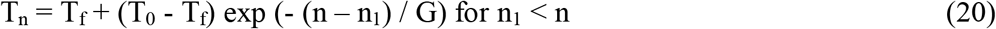

The transmission rate from a hospitalized patient is assumed to be a fraction of T_n_

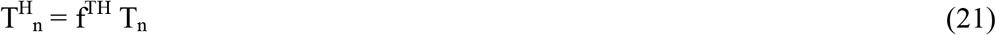

Three parameters n_1,_ G and T_f_ in equation (20) were found to be the most significant parameters affecting the number of infected cases and number of deaths from the virus. The data from countries with smaller number of cases could be fit well with smaller n_1,_ G and T_f_. In this paper, we will limit the analysis to the first outbreak of the virus in which T_n_ reduces to a final value T_f_. It may also happen that social distancing measures are lifted due to economic or social necessities before the virus is completely extinguished within the population, which would cause T_n_ to rise again from T_f_ which may lead to a recurrence. The recurrence can be modelled in future using the methodology given in this paper.

Let σ_n_ per day per person be the rate of testing of the entire population, P of the country including the infected people without symptoms (I_n_ + A_n_) on any day n. Let β_n_ per day per person be the testing rate of patients with symptoms. Except during the first few days of the virus outbreak, when testing is being developed, β_n_ = 1 in general for people with symptoms.

Cumulative number of cases with symptoms on day n is given by

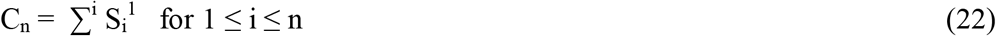

Cumulative number of reported cases which have been positively tested for the virus C^R^_n_ is given by

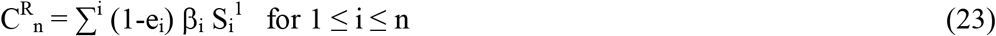

Where e_i_ is the error in reporting the number of patients on i th day. For this paper, we have assumed that the source of data [10-14] is accurate as of now, and assume e_i_ = 0 for all i. Due to imperfection in the process, often times there is error in reporting the number of cases and also the data is modified from time to time as the accounting procedure changes. The model presented here is flexible and it can incorporate changes in the data set as it is corrected from time to time. However this model is not capable of predicting suppressed case data.

Cumulative number of deaths on day n is given by

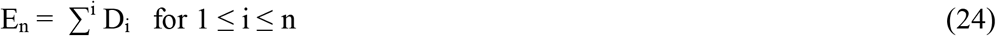

Cumulative number of reported deaths E^R^_n_ is given by

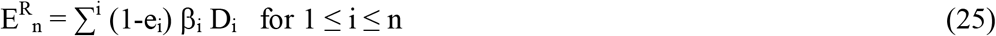

The number of active infected cases which are confirmed by testing and reported on day n can be expressed as:

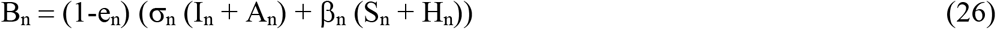

The number of reported hospitalized patients on day n is given by:

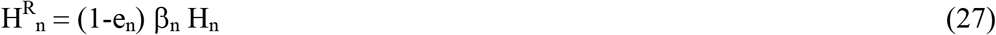

The number of reported hospitalized patients who are in ICU on day n is given by:

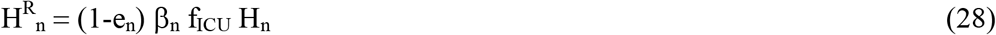

In the model, the testing rates σ_n_ and β_n_ are considered time dependent as follows:

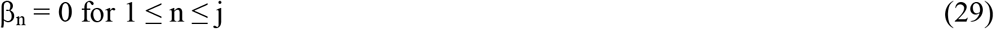

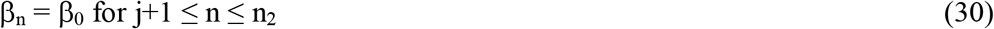

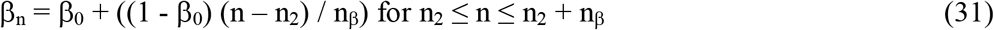

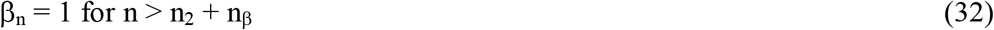

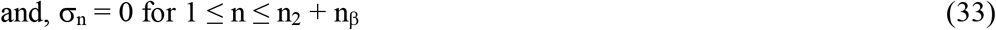

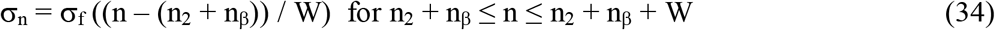

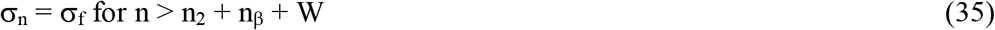

The rationale for choosing different testing rates is as follows. During the first j number of incubation days of the outbreak there is no testing and β_n_ = 0. During the first outbreak of the virus, it takes several days to identify the new virus and to develop an initial test, during which period the testing rate of the patients showing symptoms is low, denoted by β_0_. Let n_β_ be the number of days to ramp up the testing of the patients with symptoms to 100% final testing rate. The testing rate σ_n_ for the general population including those who are infected and not showing symptoms is assumed to be zero until n = n_2_ + n_β_ and let W be number of days it takes to build up the testing capability to test the general population at a final rate of σ_f_. This recognizes that large scale production of testing supplies and development of infrastructure and resources for administering of tests in large numbers take time.

### Model parameters

There are twenty one variable parameters in the model description. These are j, k, m, h, p, N_o_, T_o_, T_f_, n_1_, G, β_0_, e, n_2_, n_β_, δ_H_, δ_S_, σ_f_, W, f^TH^, f_H_ and f_ICU_. The strength of this model is that it is based on parameters which can be measured. In the beginning of the virus outbreak, the parameter values are not well known. However in the long run, the parameters values can be extracted from the actual data and the errors in model predictions can be reduced. We start with reasonable assumptions for each parameter and then use the model to fit the published data on total and daily numbers of cases and deaths for each country to iteratively optimize the values of some of the parameters.

The initial values of the parameters j, k, m, h, p, f_H_ and f_ICU_ are obtained from data presented in the literature. In this model, we have used j = 5 [18-20], k = 14 [18,20-22], m = 11 [18,20], h = 10 [18,20,22-23], p = 3 [18,21-22], q = 14 [18,20-22], f_H_ = 17% [19-20,24-25], f_ICU_ = 33% [20,24-25]. In addition we used e_i_ = 0 for all i and f_a_ = 30% [17]. The value of p varies widely due to different standards at different places.

It is to be noted that there is daily variation and probability distribution function for each of these model parameters which in principle can be measured. The time and location dependent data on these parameters for Covid-19 in each country is not available yet. When the daily values of these parameters become available on a later date, these can be inserted in the model. For each parameter such as j and m mentioned above there is a range of values. In the model, we used one constant value for each parameter which is very reasonable approximation at this stage of data availability.

We carry out multivariate optimization to fit the available data for each country. This can be considered as the training set. Starting with reasonable initial conditions in the model, iterations are made in the algorithm until the residual errors in fitting the published data for each country are minimized. At that point the final values of all parameters for this predictive model for each country become available. Once the parameter optimization is completed, the model is then run to yield information such as predicted number of cases, deaths and required hospital beds in the future. Sensitivity analysis and what if experiments can be carried out such as the effect of delay in taking initial measures. With minor modifications to the model, recurrence of the disease and response to gradual availability of a vaccine to the general population at a future date can be studied.

## References

1. Basu, S. (2020, April 28) “A Mathematical Model to Analyze Data on Coronavirus Cases” https://doi.org/10.31224/osf.io/95arg

2. Basu, S. “Modelling to Predict Hospital Bed Requirements for Covid-19 Patients in California”, medRxiv 2020.05.17.20104919; doi: https://doi.org/10.1101/2020.05.17.20104919

3. Basu, S. “Model based comparison of Covid-19 cases in two counties in the silicon valley”, medRxiv 2020.05.26.20114116; doi: https://doi.org/10.1101/2020.05.26.20114116

4. https://www.cdc.gov/coronavirus/2019-ncov/faq.html#How-COVID-19-Spreads

5. Anderson, R.M. and May, R.M., Infectious diseases of humans: dynamics and control (Oxford University Press, Oxford, 1991)

6. Hethcote, H.W. “The mathematics of infectious diseases” Society for Industrial and Applied Mathematics 42, 599–653 (2000)

7. Lin, Q., Zhao, S. et al., “A conceptual model for the coronavirus disease 2019 (COVID-19) outbreak in Wuhan, China and individual reaction and governmental action” International journal of infectious diseases. 93, 211–216 (2020)

8. Kucharski, A.J., Russell, T.W. et al., “Early dynamics of transmission and control of COVID-19: a mathematical modelling study”, www.thelancet.com/infection Published online March 11, 2020 https://doi.org/10.1016/S1473-3099(20)30144-4

9. Wu, J.T., Leung, K. and Leung, G.M., “Nowcasting and forecasting the potential domestic and international spread of the 2019-nCoV outbreak originating in Wuhan, China: a modelling study”, www.thelancet.com Vol 395, p. 689, February 29, 2020

10. https://coronavirus.data.gov.uk/#category=nations&map=rate&area=e92000001

11. https://en.wikipedia.org/wiki/COVID-19_pandemic_in_the_United_Kingdom

12. https://ourworldindata.org/covid-cases

13. https://en.wikipedia.org/wiki/COVID-19_pandemic_in_the_United_States

14. https://en.wikipedia.org/wiki/COVID-19_pandemic_in_India

15. He, X. et al., “Temporal dynamics in viral shedding and transmissibility of COVID-19”, Nature Medicine, vol 26, pp. 672–675, May 2020, www.nature.com/naturemedicine

16. https://www.cdc.gov/coronavirus/2019-ncov/hcp/clinical-guidance-management-patients.html

17. Nishiura, H., Miyama, T. et al., “Estimation of the asymptomatic ratio of novel coronavirus infections (COVID-19)”, to appear in International journal of infectious diseases (2020).

18. Chang, D. et al., “Time Kinetics of Viral Clearance and Resolution of Symptoms in Novel Coronavirus Infection”, DOI: 10.1164/rccm.202003-0524LE on March 23, 2020

19. Li, Q. et al., “Early Transmission Dynamics in Wuhan, China,of Novel Coronavirus–Infected Pneumonia”, n engl j med 382;13 nejm.org March 26, 2020

20. https://www.cdc.gov/coronavirus/2019-ncov/hcp/clinical-guidance-management-patients.html

21. Sethuraman, N., Jeremiah, S.S. and Ryo, A., “Interpreting Diagnostic Tests for SARS-CoV-2”, May 6, 2020. doi:10.1001/jama.2020.8259

22. Zhou, F. et al., “Clinical course and risk factors for mortality of adult inpatients with COVID-19 in Wuhan, China: a retrospective cohort study”, Lancet 2020; 395: 1054–62

23. https://www.cdc.gov/coronavirus/2019-ncov/hcp/disposition-hospitalized-patients.html

24. CDC COVID-19 Response Team, “Severe Outcomes Among Patients with Coronavirus Disease 2019 (COVID-19) — United States, February 12–March 16, 2020”, MMWR, vol 69(12), March 27, 2020

25. Kashyap, S., Gombar, S. et al., “Measure what matters: counts of hospitalized patients are a better metric for health system capacity planning for a reopening”, medRxiv preprint doi: https://doi.org/10.1101/2020.04.19.20072017

26. H.K. Baruah, “Nearly perfect forecasting of the total Covid-19 cases in India: a numerical approach”, medRxiv preprint doi: https://doi.org/10.1101/2020.06.13.20130096

